# Prospective validation of mean metacarpophalangeal joint extension as a measure of flexor compartment fibrotic inflammation in diabetes-related hand manifestations

**DOI:** 10.1101/2024.10.15.24315537

**Authors:** Sanat Phatak, Sarita Jadhav, Rucha Wagh, Parth Ladha, Rishi Nalkande, Rutvij Tope, Harsh Balbudhe, Rohan Shah, Smita Dhadge, Pranay Goel, Jennifer L. Ingram, Chittaranjan Yajnik

**Author notes:** Correspondence to: Dr Sanat Phatak KEM Hospital Research Centre Sardar Moodliar Road, Rasta Peth Pune 411011, India.

## Abstract

**Introduction:** Hand conditions in diabetes, namely limited joint mobility (LJM), flexor tenosynovitis (FT), carpal tunnel syndrome (CTS), and Dupuytren disease (DD), share a common pathophysiological process involving pro-fibrotic inflammation in flexor structures. A unified, quantitative measure of disease severity across these conditions is lacking, limiting correlational research. We evaluated mean metacarpophalangeal (MCP) joint extension as a potential measure of severity.

**Methods:** We assessed 2405 adults, including individuals with type 1 diabetes (n=291), type 2 diabetes (n=877), prediabetes (n=326), and non-diabetic controls (n=911). MCP extension was calculated as the average maximum passive extension of the 2nd-5th fingers, measured with a protractor. Validity was determined by correlating MCP extension with physician-rated severity (convergent) and hand grip strength and the Duruoz Hand Index (DHI, both divergent). Inter-rater reliability was tested in 128 individuals, and sensitivity to change was evaluated in 143 participants over time and in 21 rheumatology patients with subacute noninfectious FT.

**Results:** Mean MCP extension was significantly lower in individuals with all hand conditions (42.4° LJM, 42.8° FT, 39.9° DD, 51.7° CTS) than in those without (58.6°, all p<0.05). MCP extension correlated with physician-rated severity (−0.5, p<0.01) and weakly with DHI (R^2^=0.03) and grip strength (R^2^=0.07). Inter-rater reliability was strong (ICC 0.72), and MCP extension demonstrated sensitivity to change, worsening over eight months (SRM -0.61) and improving after treatment in subacute FT (SRM 0.69).

**Conclusion:** Mean MCP extension is a valid, reliable, and responsive measure for assessing fibro-inflammatory hand conditions in diabetes, suitable for use in research studies.

## Introduction

The observation that some patients with diabetes have limited joint mobility in the hand was first described by Rosenbloom et al in 1974.[1] The spectrum of hand manifestations that occur within diabetes, termed, ‘diabetic cheiroarthropathy’ or ‘diabetic hand’, now includes limited joint mobility (LJM), flexor tenosynovitis (FT) including stenosing FT or trigger finger, Dupuytren’s contracture (DC) and carpal tunnel syndrome (CTS). [2]

The clinical correlates of these hand syndromes with other organ manifestations within diabetes are unclear. While some reports show an increased prevalence of hand disorders in patients with microvascular complications of diabetes mellitus [3], others do not. [4] Despite their heterogeneity, at least three of these manifestations (LJM, FT, DC) share the common pathophysiologic process of profibrotic inflammation, demonstrated on histopathology. [5] CTS may be secondary to fibrotic thickening of the other structures in the carpal tunnel but may also be partly microvascular. [6] We previously hypothesized that these easily accessible manifestations may reflect a global profibrotic trajectory in internal organs and could potentially serve as a clinical biomarker. [7]

One hurdle in establishing such correlations is the lack of a clinical tool that can serve as a proxy measure of the amount of inflammation-fibrosis in the hand, unifying all these manifestations. The Duruoz hand index (DHI) is a functional score that has been validated for use in diabetes. [8] However, barring early FT and CTS, these conditions are rarely painful and are functionally limiting only when they produce severe mobility restriction. [9] In this situation, a clinical metric that banks on the biology of fibrosis (resistance to stretch) rather than on perceived clinical and functional effects (functional limitation, pain) is likely to be more useful. In LJM, FT and DC, profibrotic inflammation occurs in one or more of the structures on the flexor aspect of the palm. In severe cases of LJM, contractures ensue, leading to the classical ‘prayer sign’. [10] However, finger extension is likely to be limited much earlier than this state.

We hypothesized that such limitation would reflect in a reduced angle of maximum passive extension at the metacarpophalangeal (MCP) joint, when the hand is approximated on a flat surface. In a type 1 diabetes cohort, we showed that the mean of the angles of passive extension at MCP joint of the second to fifth fingers (henceforth, mean MCP extension) was reduced in those with hand manifestations. [9] Mean MCP extension is easy to calculate at the community level and does not need expensive equipment nor extensive training. Further, this metric correlated with the presence of fibrotic tissue thickening on MRI in the type 1 diabetes patients studied.

In this study, we evaluate the performance of MCP extension as a measure of hand manifestations in an expanded population that includes type 2 diabetes, prediabetes and non-diabetic controls in addition to type 1 diabetes. We use COnsensus-based Standards for the selection of health Measurement Instruments (COSMIN) methodology to describe validity, reliability and responsiveness, using separate cohorts for different aims, that add credibility to its potential use as a clinical metric of fibro-inflammatory flexor hand involvement in these conditions. [11]

## Methods

### Patients

We studied adults (>18 years) with type 1 diabetes, type 2 diabetes, prediabetes and healthy controls (Main cohort). All individuals were seen in the study period between April 2021 and April 2024 in the diabetes, internal medicine and rheumatology clinics at the King Edward Memorial Hospital, a tertiary care hospital in Pune, India. Type 2 diabetes and prediabetes were diagnosed using American Diabetes Association criteria cut-offs [12]. Type 1 diabetes was diagnosed clinically, using criteria of diagnosis prior to 30 years, documentation of ketosis and insulin dependence. Those with prediabetes and healthy controls were invited from relatives or friends accompanying patients, as well as from within the Pune Maternal Nutrition Study (PMNS) longitudinal cohort. [13] Independent recruitments were performed for inter-rater reliability and sensitivity-to change studies, and these are described under those headings (reliability cohort and responsiveness cohort, respectively).

We collected demographic and clinical information and data on diabetes duration, medications, micro-and macrovascular complications from patients’ files. Comprehensive clinical evaluations included questionnaires for hand symptoms (pain, clumsiness, finger locking, neuropathic symptoms) and a standardized examination for signs (Tinel sign, tenosynovial thickening, triggering, skin thickening, prayer sign). We measured hand grip strength using a Jamar dynamometer, (Patterson Medical, Warrenville, IL) and patients were asked to complete the DHI, a validated instrument for hand function. [8] Mean MCP extension was calculated as an average of maximum passive extension at 2nd, 3rd, 4th and 5th finger of each hand. [Figure 1A] The palm was approximated on a flat horizontal surface and the fingers were extended upwards. A protractor was used to measure the angle between the finger skin and the flat surface. All measurements were performed by trained research staff (SJ, RW, RS, RT, HB)

**Figure 1:**
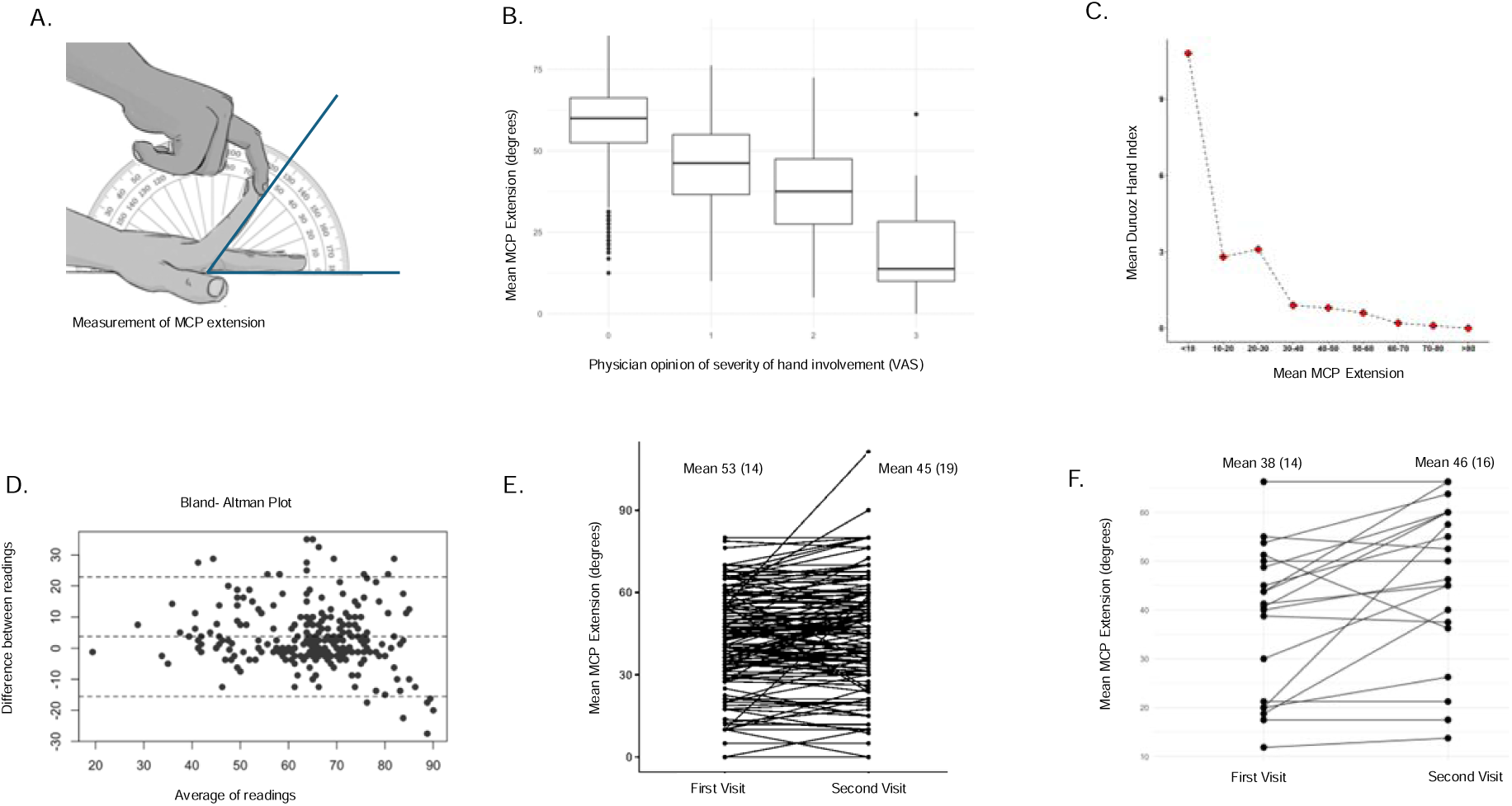
Behaviour of mean metacarpophalangeal (MCP) joint extension in diabetes-related flexor fibroinflammatory hand manifestations (diabetic hand syndromes). 1A: Clinical methodology of measuring metacarpophalangeal joint extension in the second finger; mean MCP extension is an average of the angle in the second to fifth digits. 1B: Convergent validity of mean MCP extension, correlation with physician’s opinion of severity of hand manifestations. 1C: Divergent validity of MCP extension compared to Duruoz Hand Index, a functional hand index. 1D: Bland Altman plot showing inter-rater reliability between two raters; 1E: Change in mean MCP extension in patients with hand manifestations over an average duration of eight months and 1F: Change in responsiveness to therapy for mean MCP extension in subacute flexor tenosynovitis of hands, treated with systemic or local therapies.

A rheumatologist (SP) or a diabetes specialist (SD) opined independently on the manifestation present. Interrater reliability between them (seen in 30 patients) was 0.89. LJM was diagnosed when there was skin thickening leading to flexion contractures, or a prayer sign. Those with thickened palmar skin alone but without a prayer sign were labelled as skin thickening (ST). FT was diagnosed when there was flexor tendon pain or tenderness, beading, triggering and/or palpable tendon crepitus. DD was diagnosed when palmar fascia thickening was palpable as a nodule or had resulted in a contracture. CTS was diagnosed when there was neuropathic symptoms or signs in the median nerve distribution or Tinel/ Phalen sign was positive. In addition, a physician-perceived severity of hand fibrosis-inflammation was collected on a visual analogue scale of zero to three, zero denoting no hand involvement and three being the most severely affected.

### Validity studies

#### Construct

The hand manifestations of diabetes can be considered a medical construct that results in stiffness and resistance to stretch within the flexor compartment of the hand. This condition arises from an initial period of inflammation (especially seen in subacute flexor tenosynovitis, leading to palmar pain) and later, excessive collagen deposition, leading to restricted movement. The construct focuses specifically on quantifying the mechanical properties of the affected tissues, namely their resistance to stretching forces, independent of pain perception or functional loss.

Convergent validity: We examined correlations between mean MCP extension and the physician’s subjective opinion for severity of fibro-inflammatory hand manifestations, using Spearman’s rank correlation test.

Divergent validity: We examined correlations of MCP extension with hand grip strength (measuring flexor muscle strength and nerve function) and DHI (measuring hand pain and functional loss), using Spearman’s rank correlation test.

### Reliability studies

#### Inter rater reliability

Inter rater reliability was assessed on an independent set of patients (reliability cohort) a mixture of patients with diabetes, rheumatic disease and healthy controls seen at the rheumatology and internal medicine outpatient clinics. Two examiners (PL, RN) received a brief training on measuring MCP extension. Both measured and calculated mean MCP extension for each hand, without access to the other’s measurement. Data regarding diagnosis, age and hand pain was extracted from files. We calculated single score intra-class correlation (two-way model, type-agreement) and performed Bland-Altman analysis for agreement between the two raters. Cohen’s kappa was calculated to see if both raters could pick up those with restricted mean MCP extension, taken arbitrarily as less than 40 degrees.

### Responsiveness to change

From within the main cohort, we studied individuals with at least one hand manifestation who had been examined twice, with an interval of at least 6 months. Mean MCP extension and DHI were measured at both visits. Since diabetes-related hand fibrosis is not expected to improve, we also studied a separate cohort of patients from the rheumatology clinic with subacute, painful, non-infectious flexor tenosynovitis without synovitis in the hand joints, occurring as a part of an existing autoimmune illnesses or in isolation. (Responsiveness cohort) Patients in this cohort were treated with disease modifying antirheumatic drugs (DMARDs), commonly prednisolone, methotrexate or tofacitinib or local corticosteroid infiltration as per the rheumatologist’s discretion. Mean MCP extension and DHI were measured at initial visit and after 3 months. Responsiveness to change was calculated as a Standardized Response mean (SRM) using change scores between two time points, separately for the two cohorts. Effect size was denoted with a Cohen’s D.

#### Statistical analyses

Data are presented as frequencies, mean and standard deviation as appropriate. Comparison in mean MCP extension between groups (types of diabetes vs controls, those with hand manifestations versus those without) was done using Analysis of variance (ANOVA), adjusted for age and sex. Individual statistical analyses for each goal are outlined in the respective section. All analyses were done using R. (R Core team, 2024-R Foundation for Statistical Computing, Vienna, Austria.)

#### Ethics

Ethics permission was granted by the KEM Hospital Research Centre Ethics Committee (KEMHRC/RVC/EC/1518). All patients signed informed consent forms. This study is registered with the Clinical Trials Registry of India (CTRI/2020/12/030057). All data are stored at the Diabetes Unit, KEM Hospital Research Center. All patients who were found to have hand manifestations were offered management including local glucocorticoid infiltration for trigger fingers, physical therapy and referrals to specialists (such as neurology or hand surgery, as indicated).

### Role of funding

This study is funded through a DBT/Wellcome India Alliance Clinical and Public health fellowship (IA/CPHE/19/504607, awarded to SP). The funding body had no role in study design or analysis.

## Results

### Patients’ characteristics (main cohort)

We studied 2405 participants with type 1 diabetes (n=291), type 2 diabetes (n=877), prediabetes (n=326) and non-diabetic controls (n=911). Approximately one-third of patients with diabetes and prediabetes had at least one hand manifestation present. [Table 1] LJM and ST were the most common, while isolated CTS was the rarest across patient groups. Individuals with type 1 diabetes were most likely to have multiple manifestations. Non-diabetic controls had hand manifestations, most commonly LJM and skin thickening, in 24%. Hand manifestations were more common in both type 1 diabetes, type 2 diabetes (a 10% difference) and prediabetes as compared to non-diabetic controls. (all p< 0.01)

**Table 1:**
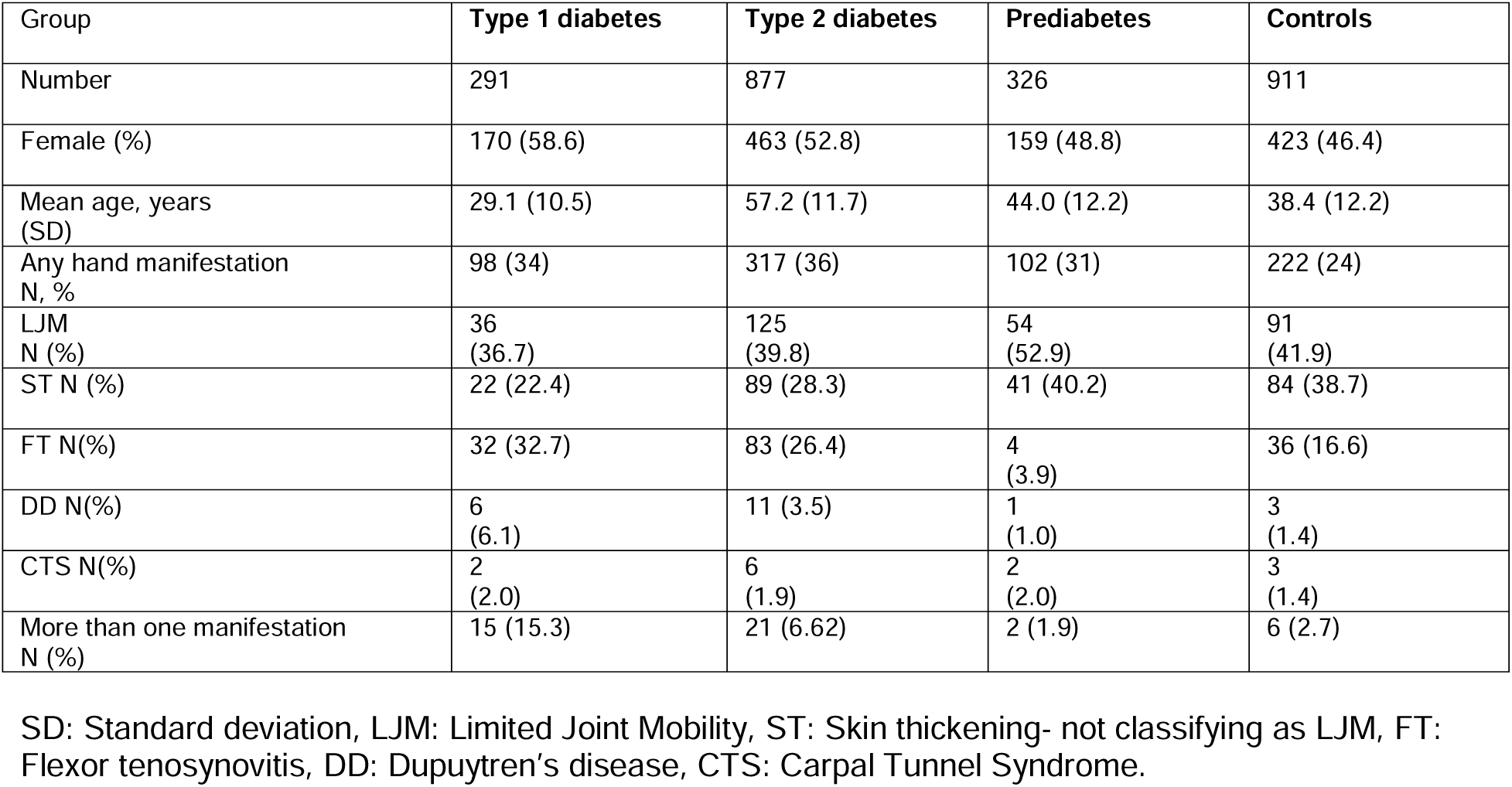
Prevalence of flexor fibro-inflammatory hand manifestations across patient groups

MCP Extension in individual manifestations and clinical associations: Mean MCP extension was lower in type 1 diabetes (mean 53.4°, p= 0.01), type 2 diabetes (mean 50.3°, p < 0.01) and prediabetes (mean 54.5°, p=0.05) than controls (mean 57.1°). Pooling all patient groups, those with no hand manifestations (n= 1666) had a mean MCP extension of 58.5°. [Table 2] Mean MCP extension was lower by approximately 20° in those with LJM, ST, FT and DD (all p<0.01) and to a lesser extent in CTS. A gender adjusted regression model in the diabetic population showed that mean MCP angle was associated with diabetes duration (p<0.01) and microvascular complications (p=0.03) but not with insulin use, body mass index, macrovascular complications, history of adhesive capsulitis or liver disease.

**Table 2:** Mean Metacarpophalangeal (MCP) joint extension and measures of hand function in patients with individual hand manifestations

|  | **None** | **LJM** | **p-value** | **ST** | **p-value** | **FT** | **p-value** | **DD** | **p-value** | **CTS** | **p-value** |
| --- | --- | --- | --- | --- | --- | --- | --- | --- | --- | --- | --- |
| **MCP extension (degrees) (mean, SD)** | 58.57<br>(11.01) | 42.48<br>(15.5) | <0.01 | 43.36<br>(12.6) | <0.01 | 42.88<br>(14.7) | <0.01 | 39.94<br>(18.4) | <0.01 | 51.73<br>(13.7) | 0.05 |
| **DHI (mean, SD)** | 0.18<br>(1.52) | 0.71<br>(4.85) | <0.01 | 0.37<br>(2.14) | 0.08 | 7.27<br>(13.49) | <0.01 | 0.42<br>(1.96) | 0.8198 | 7.23<br>(11.13) | <0.01 |
| **Grip strength (mm Hg) (mean, SD)** | 49.32<br>(19.59) | 45.13<br>(19.42) | < 0.01 | 49.83<br>(19.81) | 0.61 | 33.12<br>(14.9) | <0.01 | 54.32<br>(17.94) | 0.1057 | 38.26<br>(18.51) | 0.04 |

SD: Standard deviation, LJM: Limited Joint Mobility, ST: Skin thickening- not classifying as LJM, FT: Flexor tenosynovitis, DD: Dupuytren's disease, CTS: Carpal Tunnel Syndrome, DHI: Duruoz Hand Index

### Convergent validity

Mean MCP extension angle correlated with physician reported severity of flexor component inflammation-fibrosis (Spearman correlation coefficient -0.50, p<0.01, Figure 1B)

### Divergent validity

DHI was elevated in LJM, FT and CTS but not in ST and DD. MCP extension angle correlated weakly with DHI (R^2^ = 0.03, p<0.01), and DHI was elevated mainly in those with MCP extension below 30°. (Figure 1C) Grip strength was statistically lower in LJM, FT and CTS, and correlated weakly with MCP extension. (R^2^ = 0.07, p<0.01).

### Inter-rater reliability

The reliability cohort (n=128) included 32 patients with inflammatory or degenerative arthritis, 22 with diabetes and 2 with hypothyroidism; the rest were healthy controls. Twenty-eight had hand pain, ranging on a pain visual analogue scale from one to six. There were 256 pairs of observations for MCP extension. Two raters achieved a single score intra-class correlation of 0.72 (95% confidence intervals 0.61 to 0.79, p <0.01). On Bland Altman analysis (figure 1D), mean difference was 3.39 degrees (SD 9.49). Cohen’s Kappa for agreement in picking up those with restricted MCP extension < 40 degrees was 0.81 (p <0.01).

### Responsiveness to change

In the main cohort, 143 participants (all with at least one hand manifestation, 55 with type 1 diabetes, 82 with type 2 diabetes and 6 non-diabetes controls) had two evaluation visits at an average duration of eight months. Mean MCP extension reduced from 53 (14.6)° to 45 (19.4)° with a standardized response mean of -0.61 (p<0.01) and a Cohen’s D of 0.48. (Fig 1E) There was no significant difference in the change between type 1 and type 2 participants. Mean DHI increased from 0.77 to 3.2 (p <0.01) (data not shown).

In the smaller responsiveness cohort, we assessed 21 patients (5 (23.8%) males, mean age 56.9 years) with subacute hand flexor tenosynovitis. 11 had rheumatoid serology, four had psoriasis. Eight (38%) had type 2 diabetes, one (7.1%) had hypothyroidism, one had cicatrising conjunctivitis. Fifteen received a DMARD (methotrexate in all, tofacitinib in 12), five received local triamcinolone infiltration, all received physical therapy. After three months, mean MCP extension angle improved from 38.0 (14.8)° to 46 (16.3)°. (Fig 1F) Standardized response mean was 0.69 (p <0.01) denoting moderate sensitivity to change, with a Cohen’s D of 0.51. Mean DHI in these individuals reduced from 14.0 to 7.4 in three months, with a SRM of -0.49 (p<0.01) (data not shown).

## Discussion

Diabetes-related inflammatory-fibrotic manifestations of the hands are common. A lack of a unifying and simple methodology to measure these manifestations clinically may have precluded examining whether they are biomarkers of a global multi-organ profibrotic trajectory. We show that mean MCP extension is an easy and quantitative measurement that picks up most of these hand manifestations, even in asymptomatic stages. Using COSMIN methodology, we show that it is a valid and reliable metric on a large, heterogenous prospective cohort with varying manifestations. We lay grounds for its use in both assessing and describing the severity of hand problems themselves, as well as establishing correlations with internal organ fibrosis or serological biomarkers of ongoing profibrotic processes.

In our study, individuals with diabetes-related hand manifestations exhibited a 20-degree reduction in mean MCP angle as compared to those without, a difference we believe is clinically significant and possible to ascertain to the eye. Although grip strength also differed statistically between the groups (45 vs. 49 mm Hg in LJM versus normal), it would not be considered clinically significant. MCP extension was also reduced in LJM, skin thickening and DD, which are all painless conditions. This finding contrasted with DHI, which was not significantly higher in these three conditions and thus could pick up only CTS and FT, which affect hand function due to neuropathy or pain respectively.

The Wilson and Cleary conceptual model (1995) links biological and physiological variables to health outcomes through a continuum that includes symptom status, functional status, general health perceptions, and overall quality of life. [Figure 2] Applied to the diabetic hand, most existing scores focus on functional status or pain, assessing how patients manage daily activities or experience discomfort. To our knowledge, no instrument attempts to quantify the biology of the condition, specifically inflammation and collagen deposition, and resultant resistance to stretch. [14] Symptom status and functional status are both influenced by patient-related factors such as the patients’ pain tolerance, symptom amplification as well as environmental factors such as social and psychological support. Due to the variation that these influences would bring, they might not be good candidates for measures to correlate with other similar, biological processes.

**Figure 2:**
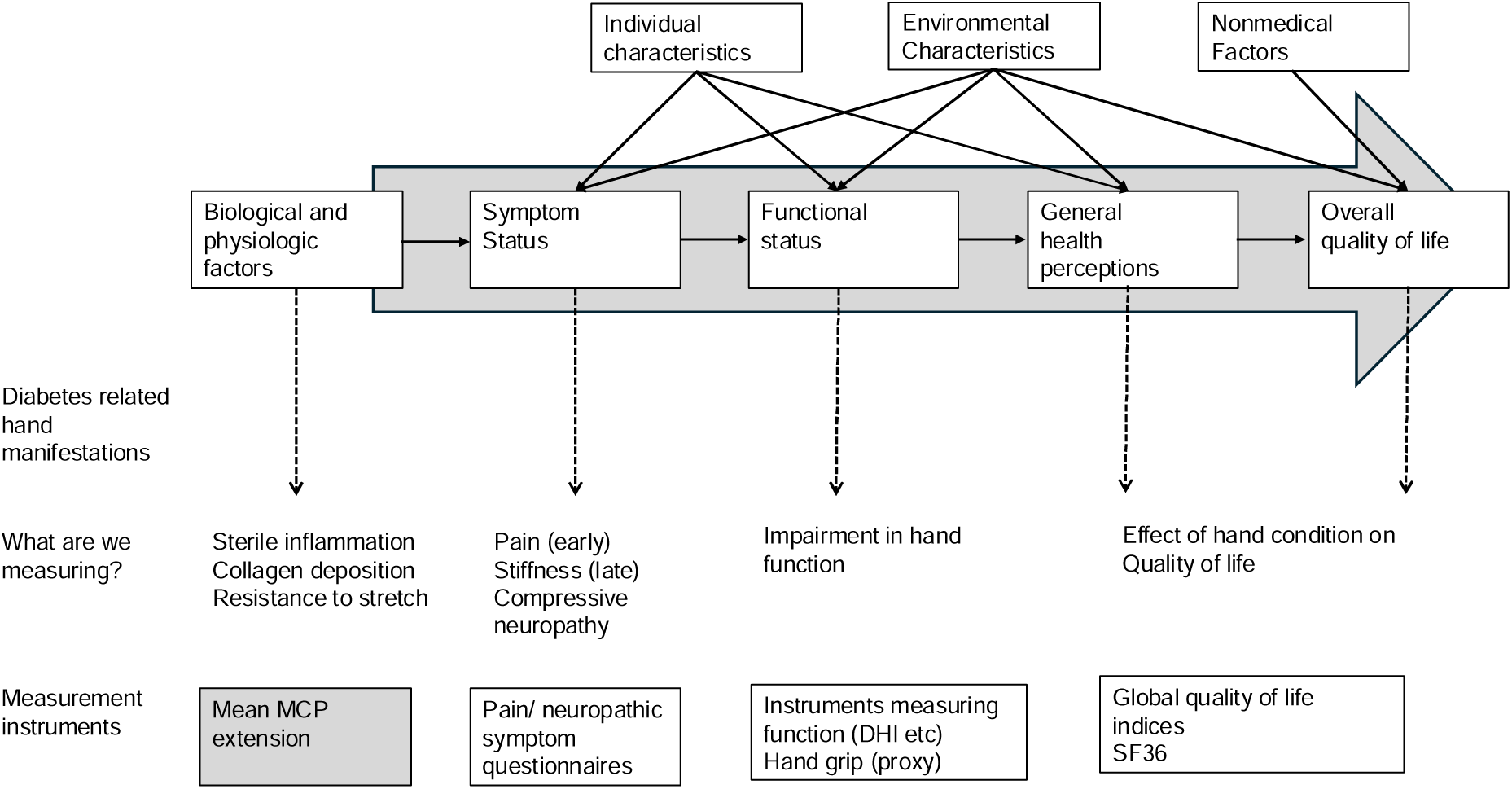
Wilson and Cleary framework (1995) to link biological factors to health-related quality of life, as applied to diabetes-related hand manifestations.

The validity data increase confidence that we are measuring the construct of interest. In this paper, we show that MCP correlated with a physician’s idea of fibrosis severity, even though this assessment is a far more subjective measure. A stronger testament to the validity was the correlation of mean MCP extension with MRI measurement of fibrous tissue deposition in flexor hand structures [9] In addition, weak correlations with both DHI and grip strength denote that these are measuring diverging constructs.

Clinically, DHI elevation in those with MCP extension <30 degrees indicate functional impairment only in the most severely affected. These data suggest that mean MCP extension adds additional value in the clinical armamentarium assessing diabetes hand.

Detecting an ongoing fibrotic process in its preclinical stages is useful because fibrosis, once established, is largely irreversible. [Henderson] Early detection may allow for timely intervention, potentially halting or slowing disease progression before irreversible tissue damage occurs. Using current diagnostic paradigms in the diabetic hand, patients will only be detected when they get the full-fledged manifestation and complain to the physician. Data suggest that end-stage phenotypes, such as the prayer sign, do correlate with stiffness in other tissues. For example, the prayer sign has been associated with difficult endotracheal intubation in surgery [15] and with increased ventilatory hours following bypass surgery [16], possibly suggesting upper respiratory and lung stiffness respectively. The prayer sign is a subjective binary sign for flexor compartment fibrosis, and such associations may benefit from quantitative measures of the same. Having a quantitative measure is also likely to help in evaluating the longitudinal course of these syndromes, which is not possible with binary outcomes. To this effect, we saw that MCP extension is sensitive to change-both in documenting a gradual decline in those with diabetes-related hand conditions and an improvement in more acute flexor tenosynovitis.

Mean MCP extension restriction before symptoms or functional deficits arise could potentially help in detecting such ‘preclinical’ disease. We used the additional category of ‘skin thickening’ as a possible preclinical state before the prayer sign is evident. Mean MCP extension was restricted in this state as compared to controls. It was also interesting to note that the group with prediabetes had a higher prevalence of ST while the fraction of LJM increased in type 1 and type 2 diabetes. This observation may suggest that the fibrotic program in the hand is set before hyperglycemia develops, which then modifies it towards increasing severity of LJM. It will be informative to follow the group with prediabetes and skin thickening, to confirm this possibility. These data will also support a Mendelian Randomization in the UK Biobank, that showed a causal relationship between hyperglycemia and hand manifestations.[17]

Reliability studies across a heterogeneous patient population increase our confidence in being able to use mean MCP extension in the clinic and community. While interrater agreement was good overall, the ability in identifying severe extension (<40 degrees) was excellent. With proper training, mean MCP extension can potentially be measured by paramedical staff in the community.

This paper has several strengths, including a large, prospective dataset of more than 2500 individuals, and the use of blinded clinical opinions to minimize bias. The inclusion of both types of diabetes, prediabetes as well as healthy controls enhances generalizability, and the use of smaller datasets tailored to the methodological approach helps establish reliability and responsiveness. The study’s weaknesses include small numbers of patients with certain hand conditions like CTS, the absence of a gold standard imaging technique, lack of correlation with similar scores like the Hand Mobility in Scleroderma (HAMIS) score [18], and the absence of long-term follow-up in most of the cohort. We did not compare this method to goniometry. Additionally, mean MCP extension is not expected to be specific to the diabetic hand or flexor tenosynovitis: synovitis as in rheumatoid arthritis, hand injuries, palmar psoriasis, neurological diseases producing contractures and stiffness are also likely to hamper MCP extension. Such populations were not evaluated in this study. Expanding the dataset to include broader hand conditions will help establish ranges. Future work will incorporate magnetic resonance imaging (MRI) as a gold standard that will add to the construct validity, as well as compare the measurement method with goniometry. Long-term follow-up studies in diabetic cheirarthropathy are few and are needed to assess the utility of detecting ‘preclinical’ hand stiffness. Finally, it would be important to correlate mean MCP extension with markers of internal organ fibrosis including the heart, kidney, lung and liver, to evaluate if the hand can indeed serve as a clinical biomarker.[7]

In conclusion, this group of studies highlights the clinical significance of mean MCP extension as a valid indicator of flexor fibro-inflammatory hand manifestations in diabetes, despite their clinical heterogeneity. A notable 20-degree difference in those with hand manifestations can be easily detected in the community. The measurement can be used reliably with minimal training and is sensitive to both worsening and improvement in these conditions. The large dataset, blinded assessments, and inclusion of controls strengthen the findings, and establish a proof-of-concept for more widespread use of this simple measure in further correlative and interventional studies.

## Data Availability

All data produced in the present study are available upon reasonable request to the authors

## Acknowledgements

We thank Dr Kalpana Jog, Dr Meenakumari (diabetes unit, KEM Hospital) and staff nurse Shivani Rangnekar for help in patient recruitment and management.

## Data Availability

Some or all datasets generated during and/or analyzed during the current study are not publicly available but are available from the corresponding author on reasonable request.

